# “There is nothing that can prevent me from supporting her:” Men’s perspectives on their involvement and support of women’s use of topical therapy for cervical precancer treatment in Kenya

**DOI:** 10.1101/2023.12.22.23300455

**Authors:** Chemtai Mungo, Konyin Adewumi, Everlyn Adoyo, Graham Zulu, Supreet Kaur Goraya, Cirillus Ogollah, Jackton Omoto, Renée M. Ferrari, Lisa Rahangdale

## Abstract

**Purpose:** Cervical cancer disproportionately impacts women in low- and middle-income countries (LMICs). The World Health Organization’s (WHO) 90/70/90 strategy aims to eliminate cervical cancer by 2030 by increasing HPV vaccination coverage to 90%, screening 70% of eligible women, and effectively treating 90% of those with abnormal results by 2030, potentially preventing 62 million deaths in LMICs. LMICs, however, struggle with limited access to cervical precancer treatment, in part due to a lack of trained professionals and weak health systems. Effective non-surgical, self-administered, which have demonstrated efficacy in high-income countries, could bridge the treatment gap in LMICs and may be more scalable and cost-effective than provider-administered therapies. To inform feasibility studies in LMICs, data are needed on the role of male partners in influencing the acceptability and uptake of self-administered topical therapies, including their support of recommended abstinence and contraception guidelines associated with these therapies.

**Methods:** Between November 2022 and April 2023, we conducted five focus group discussions (FGDs) with men aged 25 to 65 years in Kenya to explore their perspective and perceived support regarding their female partners using topical self-administered therapies for cervical precancer treatment. The FGDs were moderated by local qualitative research assistants and conducted in local languages, transcribed, coded, and analyzed using qualitative description.

**Results:** Male participants in the FGDs strongly expressed acceptance and willingness to support their wives or partners in using topical therapies for cervical precancer treatment, if available. Reported supportive behavior included permitting the use of the therapies and support of maintaining abstinence during the recommended times. Additionally, participants desired male involvement in clinic and community-based education about topical therapies to facilitate widespread support.

**Conclusion:** The use of self-administered topical therapies for cervical precancer treatment, if supported by efficacy studies in LMICs, may support achieving the WHO’s 2030 goal of 90% treatment access. We find that with adequate education, men express overwhelming support of their female partner’s use of topical therapies, including adherence to abstinence and contraception guidelines.

## 1.0 Introduction

In 2018, the World Health Organization (WHO) launched the 90/70/90 global strategy to eliminate cervical cancer, the first-ever global commitment to eliminate a cancer (1). This strategy recognizes that cervical cancer can be prevented through a combination of primary and secondary prevention and calls for 90% human papillomavirus (HPV) vaccination of all girls by age 15 years, 70% of all women receiving cervical cancer screening with a high-performance test at least twice in their lifetime, and 90% of those with an abnormal screening result adequately treated by 2030 (1). Reaching these targets would achieve an elimination threshold of 4 or fewer cases of cervical cancer per 100,000 women globally. Nowhere will these targets make a difference, as in low-and middle-income countries (LMICs), where women bear a dire and unequal burden of cervical cancer. In 2020, it is estimated that of the approximately 600,000 new cervical cancer cases and 342,000 deaths, 85% of the cases and 90% of the deaths occurred in LMICs (2). The burden of cervical cancer is particularly pronounced in sub-Saharan Africa, a reflection in part lack of established health systems (3) and the dual epidemics of human immunodeficiency virus (HIV) and HPV(4) Malawi, in Eastern Africa, has the world’s highest mortality from cervical cancer, with 51.5 deaths per 100,000 per year, twice the rate in Eastern Africa (28.6/100,000/year), and seven times the global rate (7.3/100,000/year) (5). As a result, cervical cancer is the leading cause of preventable premature cancer deaths in LMICs, accounting for 26.3% (1.83 million/6.93 million) of the total preventable premature years of life lost from cancer in 2020 (6).

While increasingly more LMICs have established HPV vaccination programs (7) and progress is being made in launching population-based screening programs, (8) access to cervical precancer treatment remains significantly limited (9–13). Cervical precancer, also known as cervical intraepithelial neoplasia (CIN), the premalignant lesion caused by persistent infection with HPV, is treatable if identified through screening, preventing progression to cervical cancer (14). However, following screening and identification of precancerous lesions, most women in LMICs lack access to treatment. In a study from Kenya, following community-based HPV screening, only 52% of those who tested HPV-positive and were referred to a health facility for treatment ultimately received treatment within six months (15). Similarly, in a retrospective study on the treatment completion following cervical cancer screening among women living with HIV in South Africa, among 2072 women with abnormal pap smears between 2013 and 2018, only 174 (25.6%) underwent guideline-indicated management within 18 months (11). Between 2011 and 2015 in Malawi, only 43.3% and 31.8% of women with precancer who required cryotherapy or excision, respectively, received treatment (16). Challenges associated with precancer treatment in LMICs include a significant loss-to-follow-up rate, as high as 40-50%, when women screened in rural facilities are required to visit central referral facilities to access treatment,(13) lack of or non-functional treatment devices at referral facilities,(16) and fragile health systems with high patient to provider ratio, resulting in significant delays in treatment access due to few or lack of trained personnel (12). In a study from the national referral hospital in Kenya, the median time to excisional treatment among those who successfully made it to the referral facility was 167 days (interquartile range 101-276 days) (17), further increasing the risk of loss-to-follow-up. To achieve the WHO target of 90% of women with cervical precancer receiving treatment globally by 2030, there is an urgent need for scalable, innovative, yet resource-appropriate strategies to close the precancer treatment gap in LMICs, including the use of patient-administered topical therapies.

While no non-surgical therapies are currently approved for the treatment of cervical precancer, the use of topical, non-excisional therapies for cervical precancer is an area of active investigation (18–24). The feasibility,(21,25,26) acceptability,(19,27) and efficacy of several topical self- or provider-administered therapies for cervical precancer treatment have been demonstrated in several studies in high-income countries (21,27–29), including randomized trials (18,19,30–32). In a randomized U.S. trial of women with cervical intraepithelial neoplasia grade 2 (CIN2), participants were randomized to 6-month observation or self-administered intravaginal 5-Fluorouracil (5FU) for primary treatment (19). Under intention-to-treat analysis, participants in the 5FU arm had a 1.62 relative risk of CIN2 disease regression (95% CI 1.10-2.56) compared to the observation arm (p=0.01), demonstrating the efficacy of self-administered 5FU cream for treating CIN2 disease. In this study, intravaginal 5FU, used once every other week for eight applications, was safe and highly acceptable, with no moderate or severe adverse events observed. In a 2020 U.S.-based single-arm Phase I study on the use of self-administered intravaginal Artesunate suppositories for primary treatment of cervical intraepithelial neoplasia grade 2 or 3 (CIN2/3), 67.9% of participants had disease regression within 15 weeks of Artesunate self-treatment (21) compared to an observed spontaneous regression rate of 28% over a similar period (33). Similarly, in this study, self-administered intravaginal Artesunate was safe and well-tolerated, with mild and self-limited adverse events. Both topical 5FU and Artesunate are on the WHO Model List of Essential Medications (34) and could feasibly be repurposed as self-administered cervical precancer treatment(19) if backed by feasibility, acceptability, and efficacy studies from LMICs. Compared to the standard-of-care provider-administered cervical precancer treatment methods, which are often inaccessible in LMICs, patient-administered topical therapies with cytotoxic or antiviral properties may be a highly scalable and cost-effective method to bridge the current precancer treatment gap in LMICs. Additionally, excisional precancer treatment methods are associated with obstetric complications, including preterm birth (35), which are particularly consequential in LMICs where access to neonatal care is limited (36).

To inform feasibility and efficacy studies on the use of topical therapies for cervical precancer treatment among women in LMICs, data are needed on the role of male partners in influencing their acceptability and uptake of such an intervention. Sexual and reproductive health (SRH) experts have long speculated about the importance of involving male partners in the SRH of women around the world (37–40). To increase women’s participation in cervical cancer screening, the WHO recommends engagement of male partners through targeted health education about cervical cancer, underscoring the crucial role of men in prevention efforts (41). Similarly, to understand the potential impact of self-administered topical therapies for cervical precancer treatment in LMICs, the role of men as decision-makers in this context should be considered (42). Recent research in Uganda and Ghana shows that, contrary to some studies that view male partners as obstacles, male partners actually support cervical cancer prevention for their wives and daughters (43–45). Such discrepancies indicate that more research is needed to understand the beliefs underlying male support of cervical cancer prevention, especially as novel treatments such as self-administered topical precancer treatments may have requirements such as contraception use or abstinence for short time frames. For many women, negotiating SRH interventions requires permission and cooperation from their male partner.

To fill this gap in the literature, we explored men’s (1) knowledge of cervical cancer, (2) perspectives on male involvement, including their intentions to support their female partner’s use of self-administered precancer therapy and the roles of male partners as facilitators to treatment uptake and adherence were they to become available for public use, and (3) beliefs about facilitators to male partner support of these therapies.

## 2.0 Methods

### 2.1 Study design & Recruitment

This cross-sectional study sequentially recruited men ages 25 to 65 years attending outpatient clinics in Kisumu County, Kenya, between November 2022 and April 2023 to participate in focus group discussions (FGDs). Inclusion criteria required that all participants have a current female partner. We used a stepped recruitment approach for the FGDs. Participants in the focus group were a subset of men who had previously participated in a survey. This survey assessed men’s views on the use of self-administered therapies for the treatment of cervical precancer in their female partners, should such treatments be recommended. All men participating in the survey were invited to participate in FGDs, but a focus was placed on recruiting men whose female partners had a history of cervical precancer treatment. A total of 39 men participated in five FGDs. A sample size of five focus groups was determined a priori based on evidence suggesting most themes are captured in three to six focus groups (46).

We adopted a constructivist paradigm to understand men’s views regarding cervical cancer screening and prevention, including the treatment of HPV and cervical precancer. We also explored their opinions, perceived acceptability, and support of their female partners’ use of self-administered topical therapies for cervical precancer treatment were it to be recommended by a health provider. Constructivism posits that understanding is derived (i.e., constructed) based on one’s perceptions, experiences, and social contexts (47). Therefore, we hypothesized that men’s acceptability of topical, self-administered therapies is based, in part, on their experiences (such as having a female partner who had ever been diagnosed with HPV or cervical precancer or cancer and prior experiences with the health system) and their social contexts (such as relationships with sexual partners).

### 2.2 Research Team

The research team included the principal investigator (CM), a Kenyan-born practicing obstetrician/gynecologist with seven years of experience, graduate students in medicine, social work, and public health (KA, SKG, GZ), a senior qualitative investigator with nearly 20 years of experience in qualitative methods and health services research (RMF), and a senior gynecologist with over 15 years experience studying topical therapies in the U.S context (LR). The focus groups were moderated and transcribed by two qualitative research assistants from the local community who spoke local languages and were conversant with the local culture (EA, JO). The moderators had training in qualitative research, prior experience conducting FDGs, familiarity with the local context, and fluency in the local languages. The moderators also received additional training from the principal investigator on the study topic, protocol, and informed consent.

### 2.3 Data Collection, Transcription, and Translation

FGDs included five to eight participants each and were held in facilities that were geographically convenient to the recruiting clinics. The FGDs were conducted in the two most spoken local languages (*Swahili* and *Dholuo*) and were guided by several domains of inquiry: 1) baseline knowledge of HPV and cervical cancer screening and prevention, 2) perception of the female partner’s risk of HPV or cervical cancer, 3) prior experience of a female partner undergoing cervical precancer treatment, 4) perceived support of and acceptability of female partners using self-administered topical therapies for HPV or cervical precancer treatment, 5) perceived barriers and facilitators of the use of topical therapies among female partners. During the FGDs, participants were introduced to two potential self-administered, intravaginal topical therapies for precancer treatment for which data are available: 5FU and Artesunate. Details provided included potential usage frequency (5FU once every other week for eight applications, Artesunate daily for five days for three cycles), abstinence requirements (two to three days of abstinence after each 5FU application and none for Artesunate), and the recommendation of consistent contraception use while using both therapies. The FGD participants’ perceptions and perceived support of their female partner’s use of topical therapies were explored in a hypothetical scenario in which the participants’ female partners needed precancer treatment and a topical therapy was recommended, with discussions about male partner support of the various requirements including abstinence or contraception requirements. Each FGD lasted approximately 90 minutes. To promote a certain degree of anonymity, participants identified themselves by respondent number (e.g., R1, R2…R8). All FGDs were audio recorded and transcribed verbatim and translated from *Swahili* and *Dholuo* to *English* by the FGD moderators. The two moderators crosschecked the translations to confirm that the translations captured all discussions that were recorded (48).

### 2.4 Data Analysis

A codebook was created during the coding process through agreement among two coders (GZ, SKG) who read and coded two of the five FGD transcripts to gain a sense of topics covered and group discussions. All FGD transcripts were coded using the developed codebook. To ensure inter-coder agreement, a subset of transcripts was randomly selected, and codes were compared for agreement; discrepancies were resolved through discussion and consensus, with revisions documented in the codebook. Content analysis was performed using NVivo V1.71.

Because the proposed topical treatment is novel in this context, our analysis involved using qualitative description, which is well-suited for increasing understanding in an area with limited knowledge (49). As this approach stays ‘close’ to the data with minimal interpretation, qualitative description supported our intent of straightforward description of participant experiences that included describing and relaying perspectives using participants’ own experiences and language. Coding reports were generated from NVivo and carefully reviewed to identify themes relating to male involvement and support of cervical precancer treatment. Themes included: 1) participants’ knowledge and awareness of cervical cancer, 2) reasons behind their intention to support their partners in the uptake of self-administered topical treatment were it to become available, 3) their perceptions of themselves as facilitators to care, and 4) education as a facilitator to male partner support.

### 2.5 Ethical Considerations

The study was approved by the ethics review boards at Maseno University School of Medicine in Kenya and the University of North Carolina at Chapel Hill in the U.S. All participants provided consent prior to participation in the study.

## 3.0 Results

Thirty-nine male participants meeting the eligibility criteria participated in five FGDs. The mean age of participants was 42.5 years (standard deviation 6.3). Most participants, 31 (79.5%), had a female partner with a history of cervical precancer treatment, 2 (5.1%) did not, and 6 (15.4%) were unsure of their female partner’s prior precancer treatment history.

### 3.1 Knowledge of cervical cancer

Most of the participants had some previous knowledge of or exposure to cervical cancer. Sources of knowledge included family or community members who had had cervical precancer or cancer, as well as television, radio, and community education sessions.

> There was a time when we were called for a brief meeting, and we were told to encourage our wives and partners to be going for cervical cancer screening and treatment. - (R4, FGD1)

Some participants mentioned having known someone who *“suffered”* from cervical cancer. One participant cited his previous exposure to cervical cancer as the driver for permitting his wife to get screening, while another cited it as motivation to learn more about the disease.

> What I have heard is that one of my relatives had her womb removed, that is hysterectomy for her to survive. She is still alive but cannot bear children, and so, whenever I hear anything to do with cancer, that memory comes to my mind and I imagine how serious the disease is. I am usually interested in knowing what type of cancer it is, and thus my curiosity to join this group and even allowed my wife to come for screening for cervical cancer. I saw my relative suffer from it and I know it is a serious thing. - (R2, FGD1)
>
> Cervical cancer is such a great problem, my sister-in-law succumbed to it…So, whenever I hear about cancer having seen my in-law suffer, I am usually keen on [learning] anything about cancer, regardless of the type. - (R3, FGD1)

Though participants had several questions about cervical cancer, and cancer in general, including questions about its acquisition, most believed that cancer was deadly, screen-able, and preventable.

> I have heard that cervical cancer, when screened or tested early, it is easy to manage, but when it has advanced, it becomes very difficult to treat. It will worsen and may not be cured. So, I usually hear that people should know their status as far as cancer is concerned early enough, so that you be put on treatment or you be sure that you do not have it. - (R8, FGD5)

### 3.2 Male partner involvement

#### 3.2.1 Intention to support female partner’s use of self-administered precancer therapy

The focus group discussions involved brief education sessions on the relationship between HPV and cervical cancer, the efficaciousness of two topical treatments currently being studied (5FU and Artesunate) as self-administered cervical precancer treatments, the treatment protocol of both drugs (including condom use and abstinence requirements for each), and the recommendation for women to use a tampon during treatment and why. When asked, all the FGD participants reported a willingness and intention to support their wife’s or female partner’s uptake of and adherence to topical, self-administered therapies for cervical precancer treatment, were it to be recommended by a healthcare provider. The beliefs underlying their intention to support were related to 1) their beliefs about cancer, 2) their understanding of the effectiveness of topical therapies presented (after it was explained to them by the FGD moderator), and 3) their perception of partner and family dynamics.

As previously mentioned, most men in the study had some baseline understanding of, or prior exposure to, cervical cancer or cancer in general. When asked if they would support their wives through cervical precancer treatment, including the use of topical, self-administered therapies, some participants stated that they would support her because they believed that cancer was deadly. However, they also believed that a cancer diagnosis was preventable if discovered and treated early. This belief was a driving factor for support.

> I can support her [to use topical self-administered therapies for precancer treatment] because cancer is a killer disease, and if it is discovered early, it can be prevented, so, I can support my wife to use the cream. - (R3, FGD3)

Men also cited believing in the effectiveness of the drugs to prevent cervical cancer as a primary reason for their acceptance and support of their use.

> I will definitely agree [to support her to use the topical therapies discussed] and she will take it too because once a disease sets in, treatment is the only remedy and cream will do it very well. - (R2, FGD5)

Men also cited their interpersonal family dynamics as reasons to support their female partner or wife’s screening and precancer treatment uptake. Some men mentioned believing that a cancer diagnosis impacted all the members of the family, including the male partner himself, while others believed that mutual support and knowing about each other’s health was an integral part of the partnership.

> There is nothing that can prevent me from supporting her, if the two of you [male and female partner] sit and talk things [through] together. - (R1, FGD4)
>
> Our wives or partners should be screened for cervical cancer so that they get treatment early enough. You see, in case our wives have cervical cancer, we as their husbands are also affected. - (R1, FGD1)
>
> Yes, I want to say that when couples live together, they live as a unit and so, they know each other’s issues and details. Even today as I came for my ARV (antiretroviral therapy) refill, she knows that I came for my drugs and so, why should she keep anything off from me, or why should I refuse that she doesn’t use the cream, yet I know it is for her own good? - (R5, FGD5)

#### 3.2.2 Male partner’s perception of themselves as facilitators to uptake and adherence

Beyond an intention to support their partner, participants believed that if they supported their partner, she would be more likely to take up and adhere to cervical precancer treatment, including the use of self-administered topical therapy. Participants emphasized that if male partners were educated about cervical cancer prevention, they would be able to present the information to their wives, positively influencing their decision to use recommended cervical precancer treatment.

Participants also emphasized the importance of their partner’s agency, stating that they would not or could not force her to use treatment, choosing to focus on support through encouragement and education.

> She will agree, after full and thorough explanation [from the male partner]. It will not be me forcing her to use the cream, but she will voluntarily accept because it is [a] source of healing. I will also take my time and explain to her the importance of this cream now that I have also understood it. I know that she will definitely agree to use it. - (R1, FGD5)

Nevertheless, most participants believed that their own (male partner) involvement would influence their female partner’s decisions to use a self-administered topical therapy, like a cream, for cervical precancer treatment.

> I would say that women usually are hesitant to use certain things because they are not sure of their husband’s reaction to whatever it is that they want to use. And so, they keep asking themselves whether the husband would be in agreement with new ideas she has learnt from elsewhere. But if there is openness in the family more so between husband and wife, then there will be free communication and the wife will use the cream fully without any fear. If the wife see[s] that you [male partner] are understanding what is going on and understanding her position, definitely she cannot refuse to use the cream. I know that men’s fear of new ideas or their failure to accept new methods would push the women into refusing to use cream. - (R8, FGD5)
>
> Once the woman is taught the importance of this cream, she will definitely use it, because as her husband, if I have come for the teaching and I now understand what it takes to use the cream and I give her my full support, she will agree to use the cream for treatment of HPV or precancer of the cervix. - (R6, FGD1)

#### 3.2.3 Ways that male partners perceive themselves to be facilitators to uptake and adherence

Participants stated ways that they intended to support their partner who may need to use self-administered therapies. Intended support highlighted ways in which men perceived themselves as facilitators of treatment uptake and adherence. This included: 1) maintaining abstinence as part of adhering to treatment protocol, 2) providing emotional support such as permission or encouragement to uptake treatment, 3) and providing financial support. Though maintaining sexual abstinence during periods of use of topical therapies has been considered a potential barrier to male partner support of topical therapies, the FGD participants unanimously asserted that they themselves were willing and able to maintain abstinence or condom use as needed if their partner were using a topical therapy, citing their understanding of the reason for the topical treatments as a key motivator of support.

> For those who have been taught like ourselves, [maintaining] abstinence is not a challenge, because we know what should be done. It will be a great challenge to those who have not been taught about cervical cancer. - (R5, FGD1)
>
> I don’t think there is any challenge in that [maintaining abstinence or using condoms]. You know the reason why you should use condoms. Also, you that there are specific times you should not have sex with your wife. If you don’t then its fine. When you see your wife applying that cream, you know why. If you know, I don’t think if there can be any problem. - (R3, FGD4)

Second to maintaining abstinence, emotional support emerged as the greatest male support-related theme throughout the FGDs. Emotional support practices cited by participants included providing their female partners permission to receive cervical cancer screening and treatment, encouraging their partner to take up treatment if they needed it, reminders to adhere to treatment timing in case of self-administered treatment used at home, maintaining open communication, and general encouragement including maintaining hope during the treatment course.

> I would allow her [to use topical therapies], and I know that she will definitely go for it [if] she was found to be positive with HPV or precancer. The use of cream is good since it will help clear the precancer and so, she will not have cancer of the cervix - (R5, FGD5)
>
> I heard it in [the] Radio station when they were announcing it. I used to hear it keenly and know what the disease is. It attacks any part of the body whether throat or private parts, so when I heard that. I did not take it seriously, it just something that is there. So later, when my wife went for screening, she [came] back and told me. They told me I had precancer, and I told her that precancer is treatable and she should [not] have stress. She became emotional and I told her that is part of life, so just be calm. You have been told you just have precancer but there are some people who have cancer. Just be calm and follow the direction that you will be given by the doctor. Later she was calm and even asked for a reassurance to know if she will be fine. She has been coming for some health talk in the hospital and she is fine now. - (R6, FGD2)

One participant noted the ways in which open communication dynamics between male and female partners could support adherence to use of self-administered topical therapies, in which case the male partner can remind his female partner to use the cream if she has forgotten.

> Just like R7 has mentioned it needs being open. Even if she forgets to apply it then you can remind her to use the cream. I don’t see any challenge in supporting them. Because if we have been supporting each other, I can’t fail to support her in that. - (R3, FGD2)

Most participants expressed a willingness to pay for the treatment, while some mentioned a willingness to pay for transportation to health facilities to receive treatment.

> I would facilitate her going to hospital like giving her fare for boda boda [bicycle] or bus fare. You also look into and arrange how the household chores that she could have executed will be performed. - (R2, FGD5)

### 3.3 Facilitators to male partner support

Male partner education emerged as the greatest facilitator of male partner support of women’s topical therapy use. Men emphasized the need for education in helping other men understand the importance of maintaining abstinence as needed when their female partner was using topical therapies.

> My wife told me about the cream but I want to learn more about it so that we, as men also get enlightened and thereby eliminating disagreement between couples. Through that [male partner education], gender-related violence will be eliminated, otherwise this thing is likely to cause chaos within marriages because the woman will want her partner or her husband to abstain because she is on medication and yet, the man doesn’t understand why he must abstain from sex for a certain period. - (R2, FGD1)
>
> The partner may resist the idea of abstinence from sex because he has not been inducted [educated] fully. The wife will tell him that they cannot have sex because she is on medication, but the husband will not understand. Therefore, it is better for men to be taught about all the information concerning cervical cancer. - (R6, FGD1)

Suggestions for male partner education included: 1) male involvement within a clinical context and 2) male partner targeted community-based education. Some participants suggested that men attend clinic visits with their partner to receive information about topical treatment from a health care provider, citing that this would make maintaining abstinence *“easier.”*

> If possible, the health care provider should tell her to come with the husband and explain things to him. So, we come together and I also listen as she is being explained to. So that is if she is given the cream then it is easier. - (R6, FGD4)

Other suggestions included providing women with educational material or *“newsletters”* from the clinic or calling male partners on the phone.

> Women go with their husbands when going to the clinic or they be given newsletters to take home for the husbands to read. - (R6, FGD3)
>
> In my view we [male partners] should just be given these forms to read. After here I will also sit down with my wife and talk about it. The best thing is to first give the women that form to take home. - (R5, FDG4)

Community-targeted education suggestions included *“group counseling”* such as having education meetings at chief barazas (meetings), providing information on the radio, providing cervical cancer treatment education in people’s homes, and male peer education.

> I would say that proper and wide information reach should be enhanced in order to include very [person] in the community. - (R3, FGD1)

Participants suggested that community-targeted education about new self-administered cervical precancer treatment could mitigate potential interpersonal conflicts or *“violence”* that may arise from the need to maintain abstinence or general ignorance about topical therapies were they to be prescribed to women.

> It can bring violence in the house, what l would request that before the use of this cream, there should be advertisements through chief barazas[meetings] or radios for us to have prior knowledge that even one day if your wife decides to do [use a self-administered treatment] it then we have a knowledge about it. - (R2, FGD3)

Another participant referenced how previous HIV/AIDs *“awareness”* campaigns that helped reduce stigma around HIV/AIDs to highlight the importance of community-targeted education methods in normalizing cervical pre-cancer treatment and increasing awareness of self-administered treatments.

> [I] am very thankful for this because it something that saves life. What we should know is how to it should be accessible to everybody. My plea is that you involve also village elders, also door to door. Distributing the health care providers as they teach people around. It [door to door teaching] can [be] easier because gathering people is also not easy. Also, there are people who are shy because they are afraid of questions that might pop in. like HIV/AIDs, when it started there were some who would not even sit here that they are hiding from people they know. For us to overcome the fear just like when HIV was here. They [healthcare workers] would come even at the house to do awareness so that people are comfortable. So, like this if it [education] can be done also in the house, where both partners are sitting together. - (R8, FGD4)

Another participant highlighted the ways in which male partners themselves could be facilitators to male partner education.

> To use [male partners] who have learnt this [cervical precancer treatment] and have been taught on this it is not hard. But for those who have [not] received the information [maintaining abstinence and using condoms] will be hard for them… We will also go outside and teach others…. There are men who will not use condoms [with] their wives. There are men who when they will see that their wife is applying that cream, it will be like there is something that woman is not telling him. We should fight this and even tell our neighbors, friends what this [topical treatment] is. Even if he [finds] - his wife using, he will say that is true. His neighbor told him. - (R7, FGD4)

## 4.0 Discussion

In this study evaluating men’s perspectives of their female partners’ use of topical, self-administered therapies for cervical precancer treatment in Kenya, we found that men were highly supportive of topical treatments after the treatment’s importance, efficacy, and use were explained to them. We also found that many participants perceived themselves to be key facilitators of their female partner’s ability and willingness to use topical self-administered therapies. All the participants reported an intention to support their partner were she to be prescribed a topical therapy. Proposed supportive behaviors included giving their partner permission to use topical therapy and adherence to abstinence and condom use requirements associated with topical therapies. Participants also highlighted the importance of male partner education, both in the clinic and in the community, to facilitate male support of topical therapies and to mitigate any potential violence resulting from not understanding the need for abstinence during treatment. Proposed education strategies included explanations about treatment from a health professional when men accompany women to the clinic, distributing educational materials to women during clinic visits to take home, and initiating community-targeted awareness campaigns to raise public awareness about topical treatments.

To our knowledge, this is the first qualitative study to explore men’s perspectives on self-administered therapies for cervical precancer treatment in an LMIC setting and their perceived support of their female partner’s use of them, should they become available. We found that men believed that male partners understanding, acceptance, and support of novel treatment methods, including self-administered therapies, would increase the likelihood of their female partner adopting and adhering to the proposed treatment. They believed that this was due, in part, to a woman’s ability to use the treatment without fear of male partner disapproval. Though they emphasized that they would not force their partners to use such a treatment, the ways in which men defined their intention to support their partners highlighted the ways that male partners may be gatekeepers to a woman’s ability to use topical, self-administered therapies (42). For instance, we found that participants viewed giving their partner permission to seek treatment as a form of support. Therefore, in settings where men are viewed as primary decision-makers of the family, a woman’s ability to use self-administered therapies for cervical precancer treatment may depend on the involvement of her male partner if she has one (42).

These findings are consistent with studies examining women’s perspectives on how male partner involvement influences their ability and likelihood of utilizing sexual and reproductive health services in LMICs (42,50–53). For example, one study found that women who attended their post-treatment follow-up visit as part of an HPV-based cervical cancer screening program in Western Kenya more often identified their male partners as supportive, compared to women who did not return and were considered “lost to follow-up” (43). This suggests that the lack of male partner support contributes to women’s inability to adhere to follow-up recommendations in this setting. In the context of self-administered therapies, a Zimbabwean study evaluating the effect of male involvement on women’s adherence to female-initiated HIV prevention methods, such as microbicide gels, found that women were more likely to use the study products if they believed their male partner supported their use (54).

Because of the pro-inflammatory nature of potential intravaginal therapies for cervical precancer, including 5FU, abstinence for specific periods around their use is recommended(18,19,32,55,56).Use of these therapies is associated with acute inflammatory changes, including transient erythema and edema of the vaginal mucosa, and mild side effects, including increased discharge, spotting, or irritation, are common(19,27,32,57). In a few cases, superficial, self-limited erosions of the vaginal or epithelial mucosa have been observed(55). Inability to adhere to abstinence recommendations while using these therapies may be associated with worse side effects, potentially increasing a woman’s risk of contracting sexually transmitted infections, including HIV(19) and may expose the partner to the agent in case of barrierless intercourse. Similarly, as some topical agents are teratogenic when used systemically, women must use contraception during their use to avoid pregnancy. Therefore, women’s ability to negotiate abstinence and contraception use with their male partners is a critical aspect of the safety and feasibility of widespread use of these therapies in contexts where women may have less agency (58). Studies generally advise two to three days of sexual abstinence following each application of 5FU and contraception throughout the treatment to prevent pregnancy.

In exploring men’s willingness to maintain abstinence for recommended periods of time when their female partners use these therapies, we found that an overwhelming majority of the FGD participants reported a willingness to maintain abstinence in support of their partner’s treatment. Participants said that following appropriate teaching or education “*abstinence or condom use is not a challenge because we know [why] it should be done*.” Participants added that maintaining abstinence may be a “*great challenge among those who have not been taught*,” emphasizing that *"as men get enlightened,"* it would help *“[eliminate] disagreement between couples*.” One participant highlighted that through male partner education around abstinence and condom use requirements, “*gender-based violence*" can be prevented; "*otherwise, this is likely to cause chaos within marriages*.” This is particularly important when condom use is recommended among married couples, as “*there are men who will not use condoms with their wives,*” and without adequate education, seeing a female partner using a topical therapy, some may feel like “*there is something [she] is not telling him*.” Though all participants claimed that they themselves could maintain abstinence, their strong emphasis on male partner “*education*” to promote treatment acceptance, especially as it pertained to abstinence and condom use, sheds light on the importance of male partner involvement in facilitating male support of topical therapies like 5FU. Despite using “*education*” as an all-encompassing term, participants’ suggestions were indicative of a desire for (1) male involvement in the treatment-seeking behaviors of their female partner and (2) awareness-raising campaigns that normalize the existence and use of topical precancer treatments. Further, participants’ emphasis on the inability of “*other men*” to maintain abstinence and the need to educate “*other men*” on abstinence requirements may be indicative of an unwillingness to acknowledge abstinence as a personal barrier to female partner support within an FGD setting.

The ability of women receiving cervical precancer treatment in LMICs to adhere to post-treatment abstinence recommendations remains a question in the literature (59). Following conventional cervical precancer treatment, guidelines recommend avoiding sexual intercourse for four to six weeks to allow healing of the cervix or using condoms for those who cannot abstain (60). Research in LMICs has found male partners to be both a perceived and experienced barrier to adherence to post-treatment abstinence recommendations (43,51). A study in Malawi assessing barriers to follow-up after an abnormal cervical cancer screening result found that although some women cited their partners as supportive of treatment, they were still unable to maintain abstinence for the recommended period (51). Another study found that women who underwent cryotherapy in Peru felt pressure from their male partners to engage in sex sooner than recommended, though this was a small minority (61). A Kenyan study found that 16% of women reported not adhering to abstinence recommendations after undergoing an excisional procedure for cervical precancer treatment (62). This discrepancy between our FGD participants’ expressed willingness to maintain abstinence was their partner to use a topical therapy requiring intermittent abstinence and women’s concerns or experiences to the contrary in the literature needs further study. It may highlight men’s preference for therapies that have shorter abstinence requirements (e.g., 2-3 days after each use of a topical therapy, compared to 4-6 weeks after an ablative or excisional procedure), providing an additional advantage of topical therapies in this context. Alternatively, the FGD participant’s responses may be affected by social desirability bias. To inform this, data are needed from LMIC-based studies on adherence to abstinence requirements among couples when a female partner is using topical therapies for cervical precancer treatment.

Suggestions for male involvement in the supporting use of topical therapies among their female partners included having male partners accompany their wives or female partners to clinic visits to seek information from healthcare professionals or sending women home with written educational materials directed towards their male partner, explaining the diagnosis and treatment requirements. This is consistent with a study in Kenya that found that male partners were willing and interested in accompanying their partner to maternal and child health to help facilitate uptake (50). Although women highly value and perceive the presence of male partners at clinical services as supportive, several SRH studies have identified obstacles to such accompaniment, including transportation costs, a need to work during clinic hours, and a lack of interest from male partners (42,53,54,63,64). Research on methods to encourage male partner participation in counseling sessions for topical precancer treatment, along with the use of education messages customized to local contexts, can shed light on how these strategies can impact the uptake of topical therapies in LMICs. Additionally, the impact of community-focused campaigns to raise awareness about possible new precancer treatments, like the topical therapies, as recommended in the FGDs, can be investigated to help normalize these treatments should they become available.

Supported by the expressed desire for male partner education, we believe that the unanimous intention to support and the general acceptance of the topical therapies reported in the FGDs was due, in part, to the information about topical therapies, including their potential use and efficacy that participants received in the focus groups. This provides further indication that medical information about topical therapies for precancer treatment, provided by a trusted “*health professional*,” is an important facilitator towards male partner involvement in the uptake and adherence to topical precancer treatment among women in LMICs.

There are several limitations to this study. Since participants were explicitly asked if they were willing to support their partner through treatment, it is possible that the responses were influenced by perceived pressure to provide a socially desirable response within a group setting. Further studies in non-group settings, including individual interviews or anonymous surveys, may further inform whether responses may differ outside a group setting. Another potential limitation is that participants in this study may not be representative of the general population due to the intentional recruitment of men whose partners had disclosed their cervical precancer treatment history, accounting for 80% of participants. Prior research suggests that an inability to seek treatment or fears about male partners as barriers to treatment often involve an inability to disclose their screening results to male partners for fears of repercussions such as accusations of promiscuity (65). Therefore, our study participants may be more likely to support topical therapies than men in the general population or those whose partners did not disclose their precancer treatment. While future studies can include a more representative sample of men, we believe that our sampling strategy is suitable for an initial study on men’s perceptions of topical therapies in this context.

In summary, in this study evaluating men’s perceptions of their female partner’s use of self-administered topical therapies for cervical precancer treatment in an LMIC, we find that, after a brief explanation of topical therapies and their potential role in precancer treatment, male study partners, all of whom had a current female partner, and a majority of whom had a partner with a history of cervical precancer treatment, were overwhelmingly accepting of their female partner’s use of self-administered topical therapies for the treatment of cervical precancer. Additionally, after receiving the said explanation, participants stated that they were willing to maintain abstinence and use condoms as necessary for treatment, though their emphasis was that "*other men*” (who may not be educated about topical therapies) may not be as willing to maintain abstinence without adequate education. Men’s perception of their influence over their partner’s ability and willingness to use such therapies highlights the ways in which male partners may be gatekeepers of their female partner’s reproductive health in this context. These findings highlight an opportunity for studies to engage male partners in ongoing and future studies investigating the use of topical therapies to help close the cervical precancer treatment gap in LMICs.

## Conflict of Interest

We declare that the research was conducted in the absence of any commercial or financial relationships that could be construed as a potential conflict of interest.

## Funding

This research was supported by the by the Eunice Kennedy Shriver National Institute of Child Health & Human Development of the National Institutes of Health under Award Number K12HD103085 and the Victoria’s Secret Global Fund for Women’s Cancers Career Development Award, in Partnership with Pelotonia Foundation and the American Association of Cancer Research (AACR). The content is solely the responsibility of the authors and does not necessarily represent the official views of the National Institutes of Health. The study funders have no role in the research.

## 11 Data Availability Statement

All relevant data is contained within the article. The original contributions presented in the study are included in the article/supplementary material. Further inquiries can be directed to the corresponding author/s.

## Notes

### Competing Interest Statement

The authors have declared no competing interest.

### Author Declarations

The study was approved by the ethics review boards at Maseno University School of Medicine in Kenya and the University of North Carolina at Chapel Hill in the U.S.

## References

1. Cervical Cancer Elimination Initiative [Internet]. [cited 2023 Nov 2]. Available from: https://www.who.int/initiatives/cervical-cancer-elimination-initiative

2. Sung H, Ferlay J, Siegel RL, Laversanne M, Soerjomataram I, Jemal A, et al. Global Cancer Statistics 2020: GLOBOCAN Estimates of Incidence and Mortality Worldwide for 36 Cancers in 185 Countries. CA Cancer J Clin. 2021 May;71(3):209–49.

3. Knaul FM, Rodriguez NM, Arreola-Ornelas H, Olson JR. Cervical cancer: lessons learned from neglected tropical diseases. Lancet Glob Health [Internet]. 2019 Mar 1 [cited 2023 Nov 26];7(3):e299–300. Available from: https://pubmed.ncbi.nlm.nih.gov/30728108/

4. Chibwesha CJ, Stringer JSA. Cervical Cancer as a Global Concern: Contributions of the Dual Epidemics of HPV and HIV. Vol. 322, JAMA. United States; 2019. p. 1558–60.

5. Gerstl S, Lee L, Nesbitt RC, Mambula C, Sugianto H, Phiri T, et al. Cervical cancer screening coverage and its related knowledge in southern Malawi. BMC Public Health. 2022;22(1):1–14.

6. Frick C, Rumgay H, Vignat J, Ginsburg O, Nolte E, Bray F, et al. Quantitative estimates of preventable and treatable deaths from 36 cancers worldwide: a population-based study. Lancet Glob Health [Internet]. 2023 Nov 1 [cited 2023 Nov 26];11(11):e1700–12. Available from: https://pubmed.ncbi.nlm.nih.gov/37774721/

7. Simms KT, Steinberg J, Caruana M, Smith MA, Lew JB, Soerjomataram I, et al. Impact of scaled up human papillomavirus vaccination and cervical screening and the potential for global elimination of cervical cancer in 181 countries, 2020-99: a modelling study. Lancet Oncol. 2019 Mar;20(3):394–407.

8. Bruni L, Serrano B, Roura E, Alemany L, Cowan M, Herrero R, et al. Cervical cancer screening programmes and age-specific coverage estimates for 202 countries and territories worldwide: a review and synthetic analysis. Lancet Glob Health [Internet]. 2022 Aug 1 [cited 2023 Nov 2];10(8):e1115–27. Available from: https://pubmed.ncbi.nlm.nih.gov/35839811/

9. Mungo, C Cohen, C, Bukusi, E Huchko M. Scaling up cervical cancer prevention in Western Kenya: Treatment access following a community-based HPV-testing approach. In: 33rd International Papillomavirus Conference (IPVC). Barcelona; 2020.

10. Castle PE, Murokora D, Perez C, Alvarez M, Quek SC, Campbell C. Treatment of cervical intraepithelial lesions. Int J Gynaecol Obstet [Internet]. 2017 Jul 1 [cited 2023 Nov 2];138 Suppl 1:20–5. Available from: https://pubmed.ncbi.nlm.nih.gov/28691333/

11. Rohner E, Mulongo M, Pasipamire T, Oberlin AM, Goeieman B, Williams S, et al. Mapping the cervical cancer screening cascade among women living with HIV in Johannesburg, South Africaa. Int J Gynaecol Obstet [Internet]. 2021 Jan 1 [cited 2023 Nov 2];152(1):53–9. Available from: https://pubmed.ncbi.nlm.nih.gov/33188707/

12. Cubie HA, Campbell C. Cervical cancer screening - The challenges of complete pathways of care in low-income countries: Focus on Malawi. Womens Health (Lond) [Internet]. 2020 [cited 2023 Nov 2];16. Available from: https://pubmed.ncbi.nlm.nih.gov/32364058/

13. Khozaim K, Orang’O E, Christoffersen-Deb A, Itsura P, Oguda J, Muliro H, et al. Successes and challenges of establishing a cervical cancer screening and treatment program in western Kenya. Int J Gynaecol Obstet [Internet]. 2014 [cited 2023 Nov 2];124(1):12–8. Available from: https://pubmed.ncbi.nlm.nih.gov/24140218/

14. Burd EM. Human papillomavirus and cervical cancer. Clin Microbiol Rev. 2003;16(1):1–17.

15. Mungo C, Ibrahim S, Bukusi EA, Truong HHM, Cohen CR, Huchko M. Scaling up cervical cancer prevention in Western Kenya: Treatment access following a community-based HPV testing approach. International Journal of Gynecology & Obstetrics [Internet]. 2021 Jan 1 [cited 2023 Nov 2];152(1):60–7. Available from: https://onlinelibrary.wiley.com/doi/full/10.1002/ijgo.13171

16. Msyamboza KP, Phiri T, Sichali W, Kwenda W, Kachale F. Cervical cancer screening uptake and challenges in Malawi from 2011 to 2015: Retrospective cohort study. BMC Public Health [Internet]. 2016;16(1):1–6. Available from: 10.1186/s12889-016-3530-y

17. Muruka K, Nelly MR, Gichuhi W, Anne-Beatrice K, Eunice CJ, Rose KJ, et al. Same day colposcopic examination and loop electrosurgical excision procedure (LEEP) presents minimal overtreatment and averts delay in treatment of cervical intraepithelial neoplasia in Kenyatta National Hospital, Kenya. Open J Obstet Gynecol [Internet]. 2013 May 8 [cited 2023 Nov 28];3(3):313–8. Available from: http://www.scirp.org/Html/2-1430361_30974.htm

18. Maiman M, Watts DH, Andersen J, Clax P, Merino M, Kendall MA. Vaginal 5-fluorouracil for high-grade cervical dysplasia in human immunodeficiency virus infection: A randomized trial. Obstetrics and Gynecology. 1999;94(6):954–61.

19. Rahangdale L, Lippmann QK, Garcia K, Budwit D, Smith JS, Le L Van. Topical 5-flourourcil for treamtent of cervical intraepithelial Neoplasia 2LJ: a Randomized Controlled Trial. The American Journal of Obstetrics & Gynecology [Internet]. 2014;210(4):314.e1–314.e8. Available from: 10.1016/j.ajog.2013.12.042

20. Desravines N, Hsu CH, Mohnot S, Sahasrabuddhe V, House M, Sauter E, et al. Feasibility of 5-fluorouracil and imiquimod for the topical treatment of cervical intraepithelial neoplasias (CIN) 2/3. Int J Gynaecol Obstet. 2023 Jul;

21. Trimble CL, Levinson K, Maldonado L, Donovan MJ, Clark KT, Fu J, et al. A first-in-human proof-of-concept trial of intravaginal artesunate to treat cervical intraepithelial neoplasia 2/3 (CIN2/3). Gynecol Oncol. 2020 Apr;157(1):188–94.

22. Fonseca BO, Possati-Resende JC, Salcedo MP, Schmeler KM, Accorsi GS, Fregnani JHTG, et al. Topical Imiquimod for the Treatment of High-Grade Squamous Intraepithelial Lesions of the Cervix: A Randomized Controlled Trial. Obstetrics and gynecology [Internet]. 2021 Jun 1 [cited 2023 Nov 5];137(6):1043–53. Available from: https://pubmed.ncbi.nlm.nih.gov/33957649/

23. Lin CT, Qiu JT, Wang CJ, Chang SD, Tang YH, Wu PJ, et al. Topical imiquimod treatment for human papillomavirus infection in patients with and without cervical/vaginal intraepithelial neoplasia. Taiwan J Obstet Gynecol. 2012;51(4):533–8.

24. De Witte CJ, Van De Sande AJM, Van Beekhuizen HJ, Koeneman MM, Kruse AJ, Gerestein CG. Imiquimod in cervical, vaginal and vulvar intraepithelial neoplasia: a review. Gynecol Oncol [Internet]. 2015 [cited 2023 Nov 5];139(2):377–84. Available from: https://pubmed.ncbi.nlm.nih.gov/26335596/

25. Desravines N, Hsu CH, Mohnot S, Sahasrabuddhe V, House M, Sauter E, et al. Feasibility of 5-fluorouracil and imiquimod for the topical treatment of cervical intraepithelial neoplasias (CIN) 2/3. Int J Gynaecol Obstet [Internet]. 2023 [cited 2023 Nov 5]; Available from: https://pubmed.ncbi.nlm.nih.gov/37431689/

26. Hampson L, Maranga IO, Masinde MS, Oliver AW, Batman G, He X, et al. A Single-Arm, Proof-Of-Concept Trial of Lopimune (Lopinavir/Ritonavir) as a Treatment for HPV-Related Pre-Invasive Cervical Disease. PLoS One [Internet]. 2016 Jan 1 [cited 2023 Nov 5];11(1). Available from: https://pubmed.ncbi.nlm.nih.gov/26824902/

27. Desravines N, Miele K, Carlson R, Chibwesha C, Rahangdale L. Topical therapies for the treatment of cervical intraepithelial neoplasia (CIN) 2-3: A narrative review. Gynecol Oncol Rep. 2020 Aug;33:100608.

28. Hendriks N, Koeneman MM, van de Sande AJM, Penders CGJ, Piek JMJ, Kooreman LFS, et al. Topical Imiquimod Treatment of High-grade Cervical Intraepithelial Neoplasia (TOPIC-3): A Nonrandomized Multicenter Study. J Immunother. 2022 Apr;45(3):180–6.

29. Desravines N, Chibwesha CJ, Rahangdale L. Low Dose 5-Fluorouracil Intravaginal Therapy for the Treatment of Cervical Intraepithelial Neoplasia 2/3: A Case Series. J Gynecol Surg. 2020;36(1):5–7.

30. Grimm C, Polterauer S, Natter C, Rahhal J, Hefler L, Tempfer CB, et al. Treatment of cervical intraepithelial neoplasia with topical imiquimod: A randomized controlled trial. Obstetrics and Gynecology. 2012;120(1):152–9.

31. Polterauer S, Reich O, Widschwendter A, Hadjari L, Bogner G, Reinthaller A, et al. Topical imiquimod compared with conization to treat cervical high-grade squamous intraepithelial lesions: Multicenter, randomized controlled trial. Gynecol Oncol [Internet]. 2022 Apr 1 [cited 2023 Nov 5];165(1):23–9. Available from: https://pubmed.ncbi.nlm.nih.gov/35177279/

32. Fonseca BO, Possati-Resende JC, Salcedo MP, Schmeler KM, Accorsi GS, Fregnani JHTG, et al. Topical Imiquimod for the Treatment of High-Grade Squamous Intraepithelial Lesions of the Cervix: A Randomized Controlled Trial. Obstetrics and Gynecology. 2021 Jun 1;137(6):1043–53.

33. Trimble CL, Piantadosi S, Gravitt P, Ronnett B, Pizer E, Elko A, et al. Spontaneous regression of high-grade cervical dysplasia: effects of human papillomavirus type and HLA phenotype. Clin Cancer Res. 2005 Jul;11(13):4717–23.

34. World Health Organization (WHO). WHO Model List of Essential Medicines - 23rd List, 2023 [Internet]. Geneva; 2023. Available from: https://www.who.int/publications/i/item/WHO-MHP-HPS-EML-2023.02

35. Liu R, Liu C, Ding X. Association between loop electrosurgical excision procedure and adverse pregnancy outcomes: a meta-analysis. J Matern Fetal Neonatal Med [Internet]. 2023 [cited 2023 Nov 5];36(1). Available from: https://pubmed.ncbi.nlm.nih.gov/36866806/

36. Narayanan I, Nsungwa-Sabiti J, Lusyati S, Rohsiswatmo R, Thomas N, Kamalarathnam CN, et al. Facility readiness in low and middle-income countries to address care of high risk/ small and sick newborns. Matern Health Neonatol Perinatol [Internet]. 2019 Dec [cited 2023 Nov 5];5(1). Available from: https://pubmed.ncbi.nlm.nih.gov/31236281/

37. Beyond II. High-Level Global Commitments Implementing the Population and Development Agenda. ICPD Beyond. 2014;

38. Wegner MN, Landry E, Wilkinson D, Tzanis J. Men as Partners in Reproductive Health: From Issues to Action. Int Perspect Sex Reprod Health [Internet]. 1998 [cited 2023 Dec 1];24. Available from: https://www.guttmacher.org/journals/ipsrh/1998/03/men-partners-reproductive-health-issues-action

39. Organization WH. Policy approaches to engaging men and boys in achieving gender equality and health equity. World Health Organization; 2010.

40. Sternberg P, Hubley J. Evaluating men’s involvement as a strategy in sexual and reproductive health promotion. Health Promot Int [Internet]. 2004 Sep 1 [cited 2023 Dec 1];19(3):389–96. Available from: 10.1093/heapro/dah312

41. Organization WH. WHO guidance note: comprehensive cervical cancer prevention and control: a healthier future for girls and women. 2013;

42. Triulzi I, Somerville C, Sangwani S, Palla I, Orlando S, Mamary HS, et al. Understanding the meanings of male partner support in the adherence to therapy among HIV-positive women: a gender analysis. Glob Health Action [Internet]. 2022 [cited 2023 Nov 20];15(1). Available from: /pmc/articles/PMC9009925/

43. Adewumi K, Oketch SY, Choi Y, Huchko MJ. Female perspectives on male involvement in a human-papillomavirus-based cervical cancer-screening program in western Kenya. BMC Womens Health [Internet]. 2019 Aug 8 [cited 2023 Nov 17];19(1):1–9. Available from: https://bmcwomenshealth.biomedcentral.com/articles/10.1186/s12905-019-0804-4

44. Binka C, Doku DT, Nyarko SH, Awusabo-Asare K. Male support for cervical cancer screening and treatment in rural Ghana. PLoS One [Internet]. 2019 Nov 1 [cited 2023 Nov 18];14(11). Available from: https://pubmed.ncbi.nlm.nih.gov/31738796/

45. de Fouw M, Stroeken Y, Niwagaba B, Musheshe M, Tusiime J, Sadayo I, et al. Involving men in cervical cancer prevention; a qualitative enquiry into male perspectives on screening and HPV vaccination in Mid-Western Uganda. PLoS One [Internet]. 2023 Jan 1 [cited 2023 Nov 18];18(1):e0280052. Available from: https://journals.plos.org/plosone/article?id=10.1371/journal.pone.0280052

46. Faulkner SL, Trotter SP. Data Saturation. The International Encyclopedia of Communication Research Methods [Internet]. 2017 Nov 7 [cited 2023 Nov 12];1–2. Available from: https://onlinelibrary.wiley.com/doi/full/10.1002/9781118901731.iecrm0060

47. Appleton J V, King L. Journeying from the philosophical contemplation of constructivism to the methodological pragmatics of health services research. J Adv Nurs. 2002;40(6):641–8.

48. Van Nes F, Abma T, Jonsson H, Deeg D. Language differences in qualitative research: is meaning lost in translation? 2010;

49. Sandelowski M. Whatever happened to qualitative description? Res Nurs Health. 2000;23(4):334–40.

50. Adegboyega A, Mollie A, Mark D, Jennifer H. Spousal support and knowledge related to cervical cancer screening: are Sub-Saharan African immigrant (SSAI) men interested?

51. Chapola J, Lee F, Bula A, Mapanje C, Phiri BR, Kamtuwange N, et al. Barriers to follow-up after an abnormal cervical cancer screening result and the role of male partners: a qualitative study. BMJ Open [Internet]. 2021 [cited 2023 Dec 1];11:49901. Available from: http://bmjopen.bmj.com/

52. Montgomery ET, Van Der Straten A, Stadler J, Hartmann M, Magazi B, Mathebula F, et al. Male partner influence on women’s HIV prevention trial participation and use of pre-exposure prophylaxis: the importance of “understanding.”

53. Nkonde H, Mukanga B, Daka V. Male partner influence on Women’s choices and utilisation of family planning services in Mufulira district, Zambia. Heliyon [Internet]. 2023 Mar 1 [cited 2023 Dec 1];9(3):e14405. Available from: /pmc/articles/PMC10025139/

54. Montgomery ET, Van Der Straten A, Chidanyika A, Chipato T, Jaffar S, Padian N. The Importance of Male Partner Involvement for Women’s Acceptability and Adherence to Female-Initiated HIV Prevention Methods in Zimbabwe. 2010;

55. Desravines N, Hsu CH, Mohnot S, Sahasrabuddhe V, House M, Sauter E, et al. Feasibility of 5-fluorouracil and imiquimod for the topical treatment of cervical intraepithelial neoplasias (CIN) 2/3. Int J Gynaecol Obstet [Internet]. 2023 Dec 1 [cited 2023 Dec 15];163(3). Available from: https://pubmed.ncbi.nlm.nih.gov/37431689/

56. Fiascone S, Vitonis AF, Feldman S. Topical 5-Fluorouracil for Women With High-Grade Vaginal Intraepithelial Neoplasia. Obstetrics and gynecology [Internet]. 2017 [cited 2023 Nov 6];130(6):1237–43. Available from: https://pubmed.ncbi.nlm.nih.gov/29112645/

57. Maiman M, Watts DH, Andersen J, Clax P, Merino M, Kendall MA. Vaginal 5-fluorouracil for high-grade cervical dysplasia in human immunodeficiency virus infection: A randomized trial. Obstetrics and Gynecology. 1999;94(6):954–61.

58. Seidu AA, Gyan Aboagye R, Okyere J, Agbemavi W, Akpeke M, Budu E, et al. Women’s autonomy in household decision-making and safer sex negotiation in sub-Saharan Africa: An analysis of data from 27 Demographic and Health Surveys. SSM-Population Health [Internet]. 2021 [cited 2023 Dec 14];14:100773. Available from: http://creativecommons.org/licenses/by/4.0/

59. Denny L, Kuhn L, De Souza M, Pollack AE, Dupree W, Wright TC. Screen-and-Treat Approaches for Cervical Cancer Prevention in Low-Resource Settings: A Randomized Controlled Trial. JAMA [Internet]. 2005 Nov 2 [cited 2023 Dec 1];294(17):2173–81. Available from: https://jamanetwork.com/journals/jama/fullarticle/201798

60. World Health Organization. World Health Organization. 2014 [cited 2023 Dec 14]. Comprehensive Cervical Cancer Control: A Guide to Essential Practice. 2nd edition. Available from: https://www.ncbi.nlm.nih.gov/books/NBK269619/

61. Coffey PS, Bingham A, Winkler JL, Bishop A, Sellors JW, Lagos G, et al. Cryotherapy Treatment for Cervical Intraepithelial Neoplasia: Women’s Experiences in Peru. J Midwifery Womens Health [Internet]. 2005 Jul 8 [cited 2023 Dec 1];50(4):335–40. Available from: https://onlinelibrary.wiley.com/doi/full/10.1016/j.jmwh.2005.04.004

62. Woo VG, Cohen CR, Bukusi EA, Huchko M. Loop Electrosurgical Excision Procedure: Safety and Tolerability Among Human Immunodeficiency Virus-Positive Kenyan Women. Obstet Gynecol. 2011;118(3):554–9.

63. Paul PL, Pandey S. An examination of the factors associated with male partner attendance in antenatal care in India. BMC Pregnancy Childbirth [Internet]. 2023 Dec 1 [cited 2023 Dec 1];23(1):1–10. Available from: https://bmcpregnancychildbirth.biomedcentral.com/articles/10.1186/s12884-023-05851-8

64. Okwako JM, Mbuthia GW, Magutah K. Barriers to male partner accompaniment and participation in maternal and child health care in Thika and Kiambu Level Five Hospital, Kenya. Pan Afr Med J [Internet]. 2023 [cited 2023 Dec 1];44(189):189. Available from:/pmc/articles/PMC10362673/

65. Bennett KF, Waller J, Ryan M, Bailey J V, Marlow LA V. Concerns about disclosing a high-risk cervical human papillomavirus (HPV) infection to a sexual partner: a systematic review and thematic synthesis. BMJ Sex Reprod Health. 2021;47(1):17–26.

